# Optimal cutoff, cognitive impairment diagnostic performance, reliability and concurrent validity of the Ascertaining Dementia 8 (AD8) questionnaire among Latinos

**DOI:** 10.1101/2025.09.19.25336192

**Authors:** Jaime Perales-Puchalt, Adam Parks, Tina Lewandowski, Jeffrey M Burns, Eric D Vidoni

**Affiliations:** Alzheimer’s Disease Research Center; Department of Neurology; University of Kansas Medical Center; Fairway, KS, USA

**Author notes:** Corresponding author: Jaime Perales-Puchalt, PhD, MPH; KU Alzheimer’s Disease Research Center; 4350 Shawnee Mission Parkway, Fairway, KS 66205.

**Keywords:** Cognitive impairment, dementia, Latinos, screening, assessment

## Abstract

The Ascertaining Dementia 8 (AD8) is a brief informant- or self-administered questionnaire designed to screen for cognitive impairment, offering several advantages over performance-based screening tests. We analyzed cross-sectional data from a non-probabilistic sample of English- and Spanish-speaking Latinos who were either cognitively unimpaired or had a research diagnosis of mild cognitive impairment or dementia. Diagnostic performance was evaluated using receiver operating characteristic (ROC) analysis, and the Youden Index was used to determine the optimal cutoff score. Internal consistency was tested with the Kuder-Richardson Formula 20, and concurrent validity with correlations to the Clinical Dementia Rating scale, Mini-Mental State Exam, and Montreal Cognitive Assessment scores. Among 46 participants, the optimal cutoff was 3 or higher for the total sample and in both language groups. At this threshold, the AD8 showed a sensitivity of 73.7% and specificity of 85.2%, with an area under the curve of 0.843. The AD8 achieved good internal consistency of 0.872 and demonstrated correlations in the expected directions with the cognitive impairment measures. The Spanish version generally outperformed the English version. The AD8 questionnaire has adequate psychometric properties and diagnostic performance among US Latinos. To our knowledge, this is the first manuscript to validate the AD8 among US Latinos. These findings support its use in healthcare settings and its applicability for multiple research purposes.

## Introduction

Dementia is a major cause of mortality and disability in later life [1] and costs the US healthcare system more than either cancer or heart disease [2]. The National Alzheimer’s Plan Act and the National Institute on Aging have identified reducing dementia disparities among ethnic minorities as a public health priority [3, 4]. Focusing on Latinos is particularly important, as they represent the fastest-growing older ethnoracial group in the US [5], are disproportionately impacted by dementia risk [6], and are projected to increase in number from 379,000 in 2012 to 3.5 million by 2060 [7].

Dementia detection rates among Latinos are lower and diagnoses are often delayed compared to non-Latino Whites [8, 9]. This detection disparity may hinder appropriate and timely treatment, which could stabilize and delay the progression of cognitive, functional, and behavioral outcomes [10, 11]. In fact, Latinos with dementia are more likely to experience neuropsychiatric symptoms [12, 13], and their relatives are more likely to experience depression than non-Latino White Americans, underscoring the need to better understand and address these detection gaps [14].

Reducing these detection disparities in dementia detection requires assessing whether instruments developed primarily with non-Latino White populations demonstrate validity and diagnostic performance when applied to Latinos. Racial and ethnic groups are characterized by distinct cultural features, including values, norms, and attitudes [15, 16]. For example, Latinos’ strong sense of caregiving duty and desire to create positive social interactions might lead to understating reports of cognitive decline [17, 18]. Latinos’ lower dementia knowledge, along with more extreme rating and acquiescent response styles might inflate reports of cognitive decline compared to other groups [19, 20]. Language is another important factor, as nearly half of older Latinos report limited English proficiency [21].

Cognitive screening tools validated among US Latinos exist, but they are limited [22–26]. Examples include commonly used tests such as the Mini Mental State Exam (MMSE), Telephone Interview for Cognitive Status (TICS), Dementia Questionnaire, and Montreal Cognitive Assessment (MoCA). These screening tools all have limitations. First, many are relatively lengthy and might not be feasible for busy primary care visits, which is where most Latinos with dementia are seen [27]. Second, most are performance tests which do not allow for proxy assessments and require a quiet room for administration. Third, they require a certain level of expertise to administer, and some require a fee for such training. Fourth, performance tests describe only the current cognitive state, but not the change from a previous state. Fifth, the only non-performance screening tool validated among Latinos, the Dementia Questionnaire, shows low diagnostic performance for cognitive impairment [26].

The Ascertaining Dementia 8 (AD8) is an ideal alternative for cognitive screening among Latinos, as it addresses the limitations listed above. The AD8 is a 3-minute yes/no self-administered questionnaire that can be completed by the informant or the patient, with less concern about noise or distractors than performance tests [28, 29]. The AD8 is simple to administer, requiring only reading the instructions and questions, and it assesses intra-individual change across a variety of cognitive domains compared to previous levels of function. This scale has shown good psychometric properties when administered to informants and patients in the US and other countries [30], including Spanish-speaking populations abroad [31–33], and at-risk populations in the US such as non-Latino Black Americans [34]. However, to our knowledge, the psychometric properties of the AD8 have not been tested among US Latinos. The present study aims to establish the optimal cutoff and test the diagnostic performance, reliability and validity of the AD8 among US Latinos.

## Methods

### Design

This was a cross-sectional secondary data analysis that uses observational data [35, 36], drawing on information provided by participants and their informants. All study procedures were approved by the Institutional Review Board of the University of Kansas Medical Center (STUDY00140406; STUDY00148941). All participants gave verbal consent for screening and written informed consent, or assent with legal representative consent for all other procedures.

### Procedures

Participants were consecutive Latino individuals interested in research opportunities who subsequently enrolled in a longitudinal observation study aligned with the National Institute on Aging National Alzheimer’s Coordinating Center (NACC). The AD8 was administered via the telephone during initial screening. Eligibility included being 65 or older (60 for understudied populations) if no memory concerns were reported, or any age if a dementia diagnosis was disclosed. Potential participants were excluded if they had a prior stroke. The observational study captured standard data, called the Uniform Data Set (UDS), at annual evaluations, which were later pooled and shared for general researcher use at NACC along with data from other centers across the country [37]. All participants received an in-depth clinical evaluation guided by the Clinical Dementia Rating (CDR) [38], a semi-structured interview of a participant and informant knowledgeable about the participant’s daily activity, and underwent a standard neuropsychological test battery and other assessments [39].

### Assessment

All data were collected in either English or Spanish, depending on the participants’ or informants’ preference. The language in which the AD8 was administered, and information about who completed the AD8, depended on the source (participant or informant). However, this latter question was not incorporated from the beginning, which led to some missing data. The survey included socio-demographic information about the participant and their informant, including age, sex, race, ethnicity, years of education, living situation, working status, and the informant’s relation to the participant.

We tested the psychometric properties of the AD8 [28]. The AD8 is available online in multiple languages, including English and an adapted Spanish version: https://otm.wustl.edu/washu-innovations/tools/ad8-licensing/. The AD8 is a brief screening tool for cognitive impairment (e.g., problems with judgment, trouble learning new things). This tool was originally developed and validated in 2005 in English in a US sample, consisting of eight yes/no questions designed to capture change from a person’s prior level of function [28, 29]. Each item is scored as 1 (Yes, a change) or 0 (No, no change), and the total score (0-8) is obtained by summing across all items. Higher scores indicate greater likelihood of dementia. Previous research has shown that AD8 scores of ≥2 were suggestive of cognitive impairment [30].

The reference standard against which the AD8 was tested for diagnostic performance was dementia or mild cognitive impairment (MCI) diagnosis. MCI and dementia diagnoses were made by a clinician or a consensus team after reviewing all available study information, with the AD8 excluded from consideration. Clinicians were instructed to consult the *Diagnostic and Statistical Manual of Mental Disorders* [40] for diagnostic purposes. In UDS version 3, all-cause dementia was determined using a modified version of the McKhann criteria, and in earlier versions, dementia was diagnosed using the standard criteria for dementia of the Alzheimer type or other non-Alzheimer dementing disorders [41, 42]. If a participant was not cognitively normal or demented, MCI in all versions was determined according to an excerpted chart from the Alzheimer’s Disease Neuroimaging Initiative (ADNI) manual based on the Petersen criteria [43]. When the clinician judged that the participant showed cognitive impairment, but the pattern, severity, or etiology did not fit research criteria for MCI or dementia, this was classified as “Impaired Not-MCI.”

We tested AD8’s concurrent validity with the English and Spanish versions of the CDR Global Score [38], and raw scores from the MMSE [44] and the MoCA.[45] These assessments were part of the Clinical Cohort, and the English and Spanish versions were the ones included in the NACC UDS [42, 46].

The CDR is a clinician-rated scale designed to assess global cognitive and functional impairment (e.g., memory, orientation, judgment). It was originally developed in 1982 in a US sample and assesses six domains: memory, orientation, judgment/problem solving, community affairs, home/hobbies, and personal care [38]. Each domain is rated on a 5-point scale, ranging from 0 (no impairment) to 3 (severe impairment), and a global score is assigned based on an algorithm considering all domains. The global score ranges from 0 (no dementia) to 3 (severe dementia). The CDR global score has demonstrated strong validity for staging dementia severity, showing good agreement with neuropathological findings [38], and with clinical diagnostic criteria for Alzheimer’s disease and related dementias [47].

The MMSE is a brief cognitive screening tool developed to screen for dementia. This tool assesses multiple cognitive domains (e.g., memory, attention, language, visuospatial ability). It was originally designed in 1975 in a US sample, with 30 items scored on a binary correct/incorrect basis [44]. The total score ranges from 0 to 30, with lower scores indicating greater cognitive impairment. The MMSE has demonstrated good reliability and validity for identifying moderate to severe dementia, but it is less sensitive to milder forms of impairment, and it shows poor specificity for executive dysfunction [48, 49].

The MoCA is a cognitive screening tool designed to detect MCI. It was originally developed in 2005 in a Canadian sample and consists of 30 items assessing multiple cognitive domains (e.g., executive function, attention, language, visuospatial skills)[45]. The total score ranges from 0 to 30, with lower scores indicating greater cognitive impairment. The MoCA has demonstrated superior sensitivity compared to the MMSE for detecting MCI and early dementia, while maintaining acceptable specificity [45, 50].

### Analysis

Data was collected prior to the regularly-scheduled NACC data freeze on August 15, 2025. Inclusion criteria in the final analytical sample required no missing data in the AD8 and reference standard diagnosis. We excluded participants categorized as Impaired-Not MCI (n=8) to improve group definition clarity. Descriptive statistics were calculated using means and standard deviations, frequencies and percentages, or medians and interquartile ranges where appropriate. We calculated concurrent validity using Spearman correlations, based on their non-parametric distribution, and used a significance level of α= 0.05 to protect against type I error. We consider correlations to be low if they range from 0.3 to 0.5; moderate if they range from 0.5 to 0.7; and high if they are higher than 0.7 [51]. We expected the AD8 to correlate with the CDR Global Score, the MMSE and the MoCA, given that these are all global cognitive constructs. We calculated internal consistency reliability using the Kuder-Richardson Formula 20 (KR-20)and the following value ranges: ≤0.5 (unacceptable), 0.7 (acceptable), 0.8 (good), and ≥0.9 (excellent) [52]. Regarding diagnostic performance, we generated receiver operating characteristic (ROC) curves in SPSS to evaluate the ability of the AD8 to discriminate MCI/dementia from unimpaired cognition. We calculated the area under the curve (AUC), sensitivity, specificity, and Youden index to identify the optimal cutoff score. We reported analyses in the total sample and stratified results by the language used to complete the AD8 (English or Spanish). We performed analyses using IBM Statistical Package for the Social Sciences (SPSS) Version 29 [53].

## Results

Table 1 summarizes the characteristics of the sample, which included 46 individuals, among which 27 were cognitively unimpaired and 19 cognitively impaired. The sample was 72.1 years old (SD=8.9) and had 9.3 years of education (SD=5.3) on average. Most (67.4%; n=31) were women and completed testing in Spanish. Among those who were cognitively impaired, 52.6% (n=10) had MCI and 47.4% (n=9) had dementia. Those with cognitive impairment tended to complete testing in Spanish, through an informant, and be younger, men, and less educated. Among the informants, 61.6% (n=8) were their adult children, 30.8% (n=4) were their spouses, 76.9% (n=10) were women, 92.3% (n=12) were Latino, and had 11.2 (SD=3.9) years of education on average.

**Table 1.** Characteristics of the total sample.

|  | Total (n=46) |  | Cognitively unimpaired (n=27) |  | Cognitively impaired (n=19) |  |
| --- | --- | --- | --- | --- | --- | --- |
|  | %/ mean<sup>a</sup>/<br>Mdn<sup>b</sup> | n/ SD<sup>a</sup>/<br>IQR<sup>b</sup> | %/ mean<sup>a</sup>/<br>Mdn<sup>b</sup> | n/ SD<sup>a</sup>/<br>IQR<sup>b</sup> | %/ mean<sup>a</sup>/<br>Mdn<sup>b</sup> | n/ SD<sup>a</sup>/<br>IQR<sup>b</sup> |
| Age <sup>a</sup> | 72.1 | 8.9 | 73.6 | 7.1 | 69.9 | 10.7 |
| Women | 67.4% | 31 | 74.1% | 20 | 57.9% | 11 |
| Tested in Spanish | 67.4% | 31 | 63.0% | 17 | 73.7% | 14 |
| Race |  |  |  |  |  |  |
| DK | 50.0% | 23 | 48.1% | 13 | 52.6% | 10 |
| White | 39.1% | 18 | 44.4% | 12 | 31.6% | 6 |
| American Indian or Native American | 2.2% | 1 | 3.7% | 1 | 0.0% | 0 |
| Other | 8.7% | 4 | 3.7% | 1 | 15.8% | 3 |
| Years of education <sup>a</sup> | 9.3 | 5.3 | 10.6 | 4.5 | 7.9 | 5.9 |
| Married | 60.0% | 27 | 65.4% | 17 | 52.6% | 10 |
| Living situation |  |  |  |  |  |  |
| With partner | 40.0% | 18 | 50.0% | 13 | 26.3% | 5 |
| Alone | 20.0% | 9 | 26.9% | 7 | 10.5% | 2 |
| With another person | 15.6% | 7 | 11.5% | 3 | 21.1% | 4 |
| With a group of people | 24.4% | 11 | 11.5% | 3 | 42.1% | 8 |
| Retired | 90.9% | 40 | 96.2% | 25 | 83.3% | 15 |
| Diagnosis |  |  |  |  |  |  |
| Unimpaired | 58.7% | 27 | 100.0% | 27 | 0.0% | 0 |
| MCI | 21.7% | 10 | 0.0% | 0 | 52.6% | 10 |
| Dementia | 19.6% | 9 | 0.0% | 0 | 47.4% | 9 |
| Completed AD8 |  |  |  |  |  |  |
| Participant | 64.9% | 24 | 80.0% | 16 | 47.1% | 8 |
| Informant | 35.1% | 13 | 20.0% | 4 | 52.9% | 9 |
| AD8 score <sup>b</sup> | 2.0 | 0.0; 4.3 | 1.0 | 0.0; 2.0 | 4.0 | 2.0; 7.0 |
| CDR Global score <sup>b</sup> | 0.0 | 0.0; 0.5 | 0.0 | 0.0; 0.0 | 0.5 | 0.5; 1.0 |
| MMSE score <sup>b</sup> | 28.0 | 24.3; 29.0 | 28.0 | 27.0; 29.5 | 22.0 | 13.0; 29.0 |
| MoCA score <sup>b</sup> | 22.0 | 16.0; 26.0 | 24.0 | 21.3; 26.8 | 16.0 | 5.0; 22.0 |
| Among those whose informant completed the AD8 |  |  |  |  |  |  |
|  | Total (n=13) |  | Cognitively unimpaired (n=4) |  | Cognitively impaired (n=9) |  |
|  | %/mean | n/SD | %/mean | n/SD | %/mean | n/SD |
| Relation |  |  |  |  |  |  |
| Children | 61.6% | 8 | 25.0% | 1 | 77.8% | 7 |
| Spouse | 30.8% | 4 | 50.0% | 2 | 22.2% | 2 |
| Friend | 7.7% | 1 | 25.0% | 1 | 0.0% | 0 |
| Women | 76.9% | 10 | 75.0% | 3 | 77.8% | 7 |
| Latino | 92.3% | 12 | 75.0% | 3 | 100.0% | 9 |
| Race |  |  |  |  |  |  |
| DK | 61.5% | 8 | 25.0% | 1 | 75.0% | 7 |
| White | 38.5% | 5 | 75.0% | 3 | 22.2% | 2 |
| Years of education<sup>a</sup> | 11.2 | 3.9 | 15.0 | 3.8 | 9.6 | 2.7 |

a: Mean and standard deviation (SD) used due to normal distribution; b: Median (Mdn) and interquartile range (IQR) used due to non-normal distribution; DK: Don't know; CDR Global: Clinical Dementia Rating Global score; MMSE: Mini Mental State Exam raw score; MoCA: Montreal Cognitive Assessment raw score.

Shown in Table 2 are the AD8 diagnostic performance statistics for the AD8. Sensitivity and specificity values were derived from the coordinates of the ROC curve. The Youden’s index was highest for cutoff scores of 3 for the total sample, and for those tested in Spanish and English. For a cutoff of 3, the sensitivity was 0.737 and specificity was 0.852 for the total sample. At this level, the sensitivity was higher for the Spanish group (0.786) and specificity was higher for the English speakers (0.900). The AUC for the total sample was 0.843 (Standard Error= 0.063), and the KR-20 reliability index was 0.872, indicating good reliability. These estimates performed better among those tested in Spanish compared to those in English (see Figure 1).

**Table 2.** Sensitivity and Specificity of the AD8 in detecting cognitive impairment in the Total, ^a^ Spanish and English-speaking groups ^b^.

| Score | Sensitivity |  |  | Specificity |  |  | Youden's Index |  |  |
| --- | --- | --- | --- | --- | --- | --- | --- | --- | --- |
|  | Total | Spanis<br>h | Englis<br>h | Total | Spanis<br>h | Englis<br>h | Total | Spanis<br>h | Englis<br>h |
| 1 | 0.895 | 0.929 | 0.800 | 0.481 | 0.412 | 0.600 | 0.376 | 0.340 | 0.400 |
| 2 | 0.842 | 0.929 | 0.600 | 0.704 | 0.647 | 0.800 | 0.546 | 0.576 | 0.400 |
| 3 | **0.737** | **0.786** | **0.600** | **0.852** | **0.824** | **0.900** | **0.589** | **0.609** | **0.500** |
| 4 | 0.579 | 0.714 | 0.200 | 0.889 | 0.882 | 0.900 | 0.468 | 0.597 | 0.100 |
| 5 | 0.474 | 0.571 | - | 0.926 | 0.941 | - | 0.400 | 0.513 | - |
| 6 | 0.421 | 0.571 | 0.000 | 1.000 | 1.000 | 1.000 | 0.421 | 0.571 | 0.000 |
| 7 | 0.316 | 0.429 | - | 1.000 | 1.000 | - | 0.316 | 0.429 | - |
| 8 | 0.211 | 0.286 | - | 1.000 | 1.000 | - | 0.211 | 0.286 | - |

Bold: highest Youden's Index indicating optimal cutoff score.

**Figure 1.**
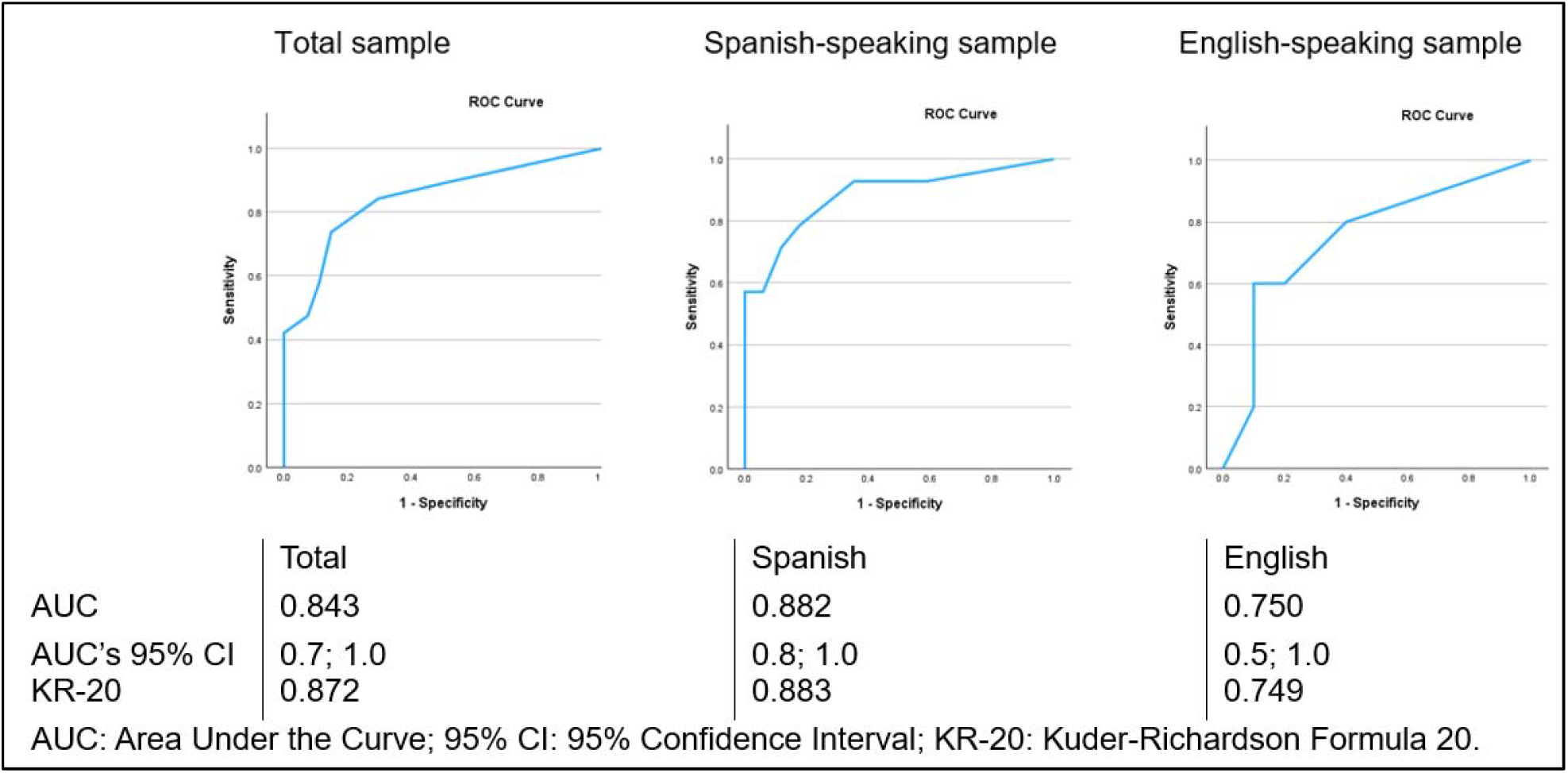
Receiver operating characteristic curves, area under the curve and internal consistency for the AD8 for cognitive impairment (diagnosis of dementia or MCI) vs cognitively unimpaired

As presented in Table 3, the AD8’s concurrent validity was evaluated by Spearman correlations with the CDR Global, MMSE, and MoCA scores. All correlations were in the expected direction: the higher the AD8 score, the higher the CDR Global score and the lower the MMSE and the MoCA scores. In the total sample, the continuous AD8 score had a spearman correlation of 0.687 with the CDR global, −0.378 with the MMSE and −0.388 with the MoCA (p<0.05 for all). Correlation trends persisted across languages and when using the discrete AD8 score of 3 or higher. These were moderate and statistically significant for the CDR global and small-moderate, yet non-statistically significant for the MMSE and MoCA scores.

**Table 3.** Spearman correlations between AD8 continuous or discrete score (≥3) and cognitive variables.

|  | Total |  | Spanish |  | English |  |
| --- | --- | --- | --- | --- | --- | --- |
| AD8 as continuous score |  |  |  |  |  |  |
|  | Correlation | p value | Correlation | p value | Correlation | p value |
| CDR Global | **0.687** | **<0.001** | **0.717** | **<0.001** | **0.611** | **0.016** |
| MMSE | **-0.378** | **0.016** | -0.316 | 0.108 | -0.475 | 0.101 |
| MoCA | **-0.388** | **0.015** | -0.372 | 0.056 | -0.215 | 0.502 |
| AD8 as discrete score (≥3) |  |  |  |  |  |  |
| CDR Global | **0.649** | **<0.001** | **0.634** | **<0.01** | **0.659** | **0.008** |
| MMSE | -0.188 | 0.245 | -0.188 | 0.347 | -0.257 | 0.396 |
| MoCA | -0.269 | 0.098 | -0.247 | 0.215 | -0.451 | 0.141 |

CDR Global: Clinical Dementia Rating Global score; MMSE: Mini Mental State Exam raw score; MoCA: Montreal Cognitive Assessment raw score

## Discussion

The current manuscript aimed to test the diagnostic performance, reliability and validity of the AD8 among US Latinos. To our knowledge, this is the first test of the psychometric properties of the AD8 in this population. We found that a threshold of 3 or higher has the highest diagnostic performance. We also found that the AD8 demonstrates good reliability and concurrent validity, meaning that it measures what it is supposed to measure (cognitive impairment) with adequate accuracy. The AD8 performed particularly well among Spanish speakers.

The results showing an optimal AD8 cutoff of 3 or higher for identifying combined cognitive impairment and dementia contrasts with the original largely non-Latino White American validation studies [29]. In those studies, the strongest Youden indices were observed at thresholds of 1 or higher for the patient version and 2 or higher for the informant version. Cutoff scores have also varied across cultures. For example, in Spain, the optimal cutoff score was found to be 4 or higher for the informant version [31], while a Korean study found that the optimal cutoff was 3 or higher with the patient version [54]. These differences underscore the importance of testing diagnostic performance in different populations, although they may also reflect differences in administration method (informant vs. patient) and the distribution of MCI or dementia within diagnostic groups.

In this Latino sample, the AD8 achieved an AUC of 0.843 overall and 0.882 among Spanish speakers. Compared with other screening tools tested among Latinos, the AD8’s performed similarly to the TICS and the picture version of the Free and Cued Selective Reminding Test with Immediate Recall (AUC=0.86 and 0.86) [26, 55]. The AD8 showed stronger diagnostic performance than the MMSE and MoCA (AUCs=0.72 and 0.79, respectively) [25, 55], and the questionnaire-based Dementia Questionnaire (AUC=0.71) among Latinos [26]. The AD8 in our Latino sample also achieved a good internal consistency (0.872), and correlated as expected with other cognitive scales, namely the CDR Global (0.687), MMSE (−0.378) and MoCA scores (−0.388). These findings are consistent with the US English validation of the AD8, which reported internal consistency of 0.86 for the informant version [56], correlations with the CDR Global scores of 0.76 (informant) and 0.36 (patient), and with the MMSE scores of −0.53 and −0.27, respectively [29]. They also align with Spain’s AD8 validation, which showed internal consistency of 0.90 and a correlation of 0.72 with the CDR Global score for the informant version [31]. Together, these findings suggest that the AD8 is superior to other questionnaire-based cognitive screening tools, and comparable, if not superior, to more resource-intensive performance-based tools among Latinos.

Limitations of this study include the small sample size, reliance on both informant and participant-completed assessments (with participant versions typically less robust), and the use of a non-probabilistic sample that limits generalizability. In addition, the reference standard did not differentiate MCI from dementia, though both were included. Finally, the small number of English-speaking participants limited subgroup analyses and may have contributed to reduced psychometric performance in this group.

Despite, these limitations, this manuscript has implications for public health and research. This manuscript provides the first validation of the AD8 among US Latinos. The AD8 can now be used more accurately in clinical and research settings with a cutoff of 3 or higher compared to the commonly cited cutoff of 2 or higher for the Spanish version [57], based on a US English-speaking, non-Latino White sample [28]. Clinicians can utilize the AD8 in busy primary care clinics, the first and often only point of healthcare contact for many Latinos [27], knowing it has sound psychometric properties. The tool’s brevity, flexibility of administration to individuals (patient or informant) and in methodology (in-person or by telephone), reduced sensitivity to distractions, and ease of use by minimally trained staff make it especially practical. These advantages also extend to research, where the AD8 could be incorporated into eligibility screening, intervention protocols, or outcome measures.

Future studies should examine the psychometric properties of informant- and participant-completed versions of the AD8 separately in larger Latino samples. Studies should also use probabilistic sampling methods, distinguish MCI from dementia in reference standards, and stratify by country of Latino descent to assess cultural generalizability. Evaluating predictive validity through associations with longitudinal cognitive change and neuroimaging outcomes will also be important. Given that the US Preventive Services Task Force has not yet recommended for or against universal cognitive screening due to insufficient evidence [58], the AD8 could serve as a valuable tool for testing whether systematic screening improves diagnosis, treatment, and care among Latinos.

As the number of Latinos with dementia continues to grow, valid and reliable diagnosis of cognitive impairment will be critical. The need affects both research, where Latinos are underrepresented, and clinical practice, which too often relies on a one-size-fits-all approach. This study provides the first validation of the AD8 in US Latinos. Its sound psychometric properties among both English- and Spanish-speaking Latinos support its use in busy clinic settings and for multiple research purposes. Future studies should build upon this foundation to strengthen the evidence for this promising diagnostic tool.

## Data Availability

Data produced in the present study are available upon reasonable request to the authors.

## Acknowledgements

Authors are thankful to JUNTOS, Don Bosco, Guadalupe Centers, the Landon Center on Aging, Duchesne and Vibrant Health clinics, El Centro inc. UsAgainstAlzheimer’s, the GAP Foundation, the Alzheimer’s Association, and the Community Partnership for Health for their help building our community network.

## Statements and Declarations; Compliance with Ethical Standards

- Disclosure of potential conflicts of interest: The authors have no relevant financial or non-financial interests to disclose.
- Research involving Human Participants and/or Animals: All study procedures were approved by the Institutional Review Board of the University of Kansas Medical Center (STUDY00140406; STUDY00148941).
- Informed consent: All participants gave verbal consent for screening and written informed consent or assent with legal representative consent for all other procedures.

## Data availability statement

De-identified data are available from the corresponding author upon reasonable request and subject to approval.

## Author contributions

Conceptualization: JPP; Methodology: All authors; Formal analysis and investigation: JPP; Writing - original draft preparation: JPP; Writing - review and editing: All authors; Funding acquisition: JMB and EDV; Resources: All authors.

